# The impact of the termination of Lymphatic Filariasis mass drug administration on Soil-transmitted Helminth prevalence in school children in Malawi

**DOI:** 10.1101/2024.10.21.24315853

**Authors:** Faduma Farah, Claudio Fronterre, Mark Taylor, Armelle Forrer

## Abstract

**Background:** Soil-transmitted helminths (STH) have been passively treated with the implementation of mass drug administration (MDA), with the drugs ivermectin and albendazole, against the parasitic disease, lymphatic filariasis (LF). In Malawi, LF MDA was administered to communities between 2008 and 2014. The aim of this analysis is to estimate the impact of LF MDA and its termination on STH prevalence in school aged children.

**Methodology:** School survey data of STH prevalence in Malawi were obtained through the ESPEN website. The surveys spanned the periods before (1998-2004), during (2012-2014) and after LF MDA (2015-2019). Bayesian mixed-effects models were fit to estimate the impact of LF MDA termination, and other STH risk factors, on the odds of infection as well as generate predictions of nationwide STH prevalence after LF MDA.

**Principal findings:** School children after the termination of LF MDA had a threefold increase in the odds of *A. lumbricoides* infection compared to school children during the implementation of LF MDA (Odds Ratio (OR): 3.4, 95% credible interval (CI): 1.99 – 5.94), despite ongoing STH preventive chemotherapy targeting school age children. In contrast, school children had lower odds of hookworm infection after LF MDA compared to during (OR: 0.5, 95% CI: 0.33 – 0.73). Mulanje district in the south has above 50% probability of exceeding 20% *A. lumbricoides* prevalence while the probability for exceeding 20% hookworm prevalence is below 50% nationwide.

**Conclusions/significance:** An overall resurgence in *A. lumbricoides* infections after LF MDA is identified in school children despite ongoing annual STH preventive chemotherapy. Monitoring of STH prevalence and infection intensity using high sensitivity diagnostics should be prioritised to surveil this resurgence and better delineate infection hotspots. A greater assessment of underlying factors would also further aid the identification of hotspots.

**Author Summary:** Soil-transmitted helminths (STH) are a group of parasitic worms that cause infections. They cause significant morbidity in children and women and the World Health Organisation (WHO) recommends the mass distribution of drugs to these populations to treat infections. In Malawi, distribution of albendazole to school age children to treat STH has been conducted annually since at least 2012. In the past, Malawi also distributed albendazole to whole communities to treat another parasitic disease, lymphatic filariasis (LF). The aim of this study was to investigate the impact of terminating LF treatment on STH prevalence in school children. We found that after mass LF treatment was stopped, school children had around three times the odds of infection for one species of STH, compared to school children during the time of LF treatment distribution. This indicates a resurgence in infection, despite ongoing STH treatment, after community-wide treatment against LF was stopped. To better understand the lack of community treatment and its impact, monitoring of STH prevalence and infection intensity with more sensitive diagnostics needs to be prioritised to avoid further resurgence of infection. An understanding of underlying factors, such as population movement or the potential emergence of drug resistance, would also help in identifying hotspots.

## Introduction

Soil-transmitted helminths (STH) are parasitic worm infections that predominately burden tropical countries of low-income with limited access to adequate sanitation. They are classed as a neglected tropical disease (NTD) and are estimated to infect 1.5 billion people globally (1). In 2019, the burden was estimated to be equivalent to 1.9 million disability-adjusted life years (2). The main STH species are *Ascaris lumbricoides*, *Trichuris trichiura* and the hookworms *Ancylostoma duodenale* and *Necator americanus*. Infection occurs through either ingestion of the helminth eggs (*A. lumbricoides* and *T. trichura*) or direct skin penetration of larvae (hookworms), which are both acquired from faecal contaminated soil. Chronic infections can lead to malnutrition, iron-deficiency anaemia and stunting of child development, making women of reproductive age and children at significant risk of morbidity (3, 4). The prevalence of infection with either *A. lumbricoides* or *T. trichiura* peaks in children 5-15 years of age (3), where with hookworm infections, prevalence increases with age and peaks in adulthood (5).

The World Health Organisation (WHO) has targeted STH for elimination as a public health problem, which is classified as a < 2% prevalence of moderate to heavy intensity (M&HI) infections (6). Preventive chemotherapy (PC) programmes with the anthelmintic drugs, albendazole or mebendazole, target countries where prevalence exceeds 20%. STH PC is targeted primarily at school aged children through school distributions. Pre-school aged children and women of reproductive age are also target groups for treatment. However, treatment coverage for pre-school aged children is highly variable and only a few countries are known to actively treat women of reproductive age (7, 8). Re-infection rates between treatment rounds are high where adequate water and sanitation infrastructure is not in place (9), which threatens the attainability of the WHO goals and questions the current method of targeted deworming that is employed.

An alternative to targeted deworming of school children is treatment of whole communities at risk. Modelling studies have shown that elimination of STH or achievement of the WHO goals need community wide mass drug administration (MDA) (10, 11). As a result, trials are currently underway to determine the ability of community wide MDA to interrupt transmission of STH (12). An example of community-based MDA that passively treats STH infections is MDA against the parasitic disease lymphatic filariasis (LF). This involves the distribution of albendazole in combination with either ivermectin, where countries are also endemic for the disease onchocerciasis, or diethylcarbamazine. With LF MDA currently implemented in 45 countries and 18 having completed the required number of rounds, effort is being made to understand how to sustain the impact of this on STH prevalence (13–16). In Malawi, STH PC has been implemented nationwide since at least 2012 (17). LF MDA was implemented in Malawi, alongside STH PC, from 2008 to 2014, when it was then terminated. Following successful transmission assessment surveys, the country was certified by the WHO as having eliminated LF as a public health problem in 2020 (18).

The aim of this study is to analyse the impact of LF MDA termination on STH prevalence in school children, map STH prevalence before, during and after LF MDA, and predict STH prevalence post-termination of LF MDA across Malawi. Risk factors for STH infection and predictions of prevalence in Malawi post LF MDA are determined using Bayesian models of open data on STH prevalence in school children, available through the Expanded Special Project for Elimination of Neglected Tropical Diseases (ESPEN) website. The potential presence of spatial correlation in the data was also checked. The predictions will assess whether STH have persisted despite several years of community-based albendazole treatment and identify any potential hotspots for infection.

## Methods

### Study area

Malawi is a landlocked country in Sub-Saharan Africa, bordered by Mozambique, Tanzania and Zambia. Malawi has a tropical climate with the rainy season between November and April. The country contains varying altitude, with mountainous areas as well as low lying regions in the south of around 90m above sea level. Malawi’s population is estimated at 20.5 million in 2022, of which 43% are children (0-14 years). Around 70% of the population live under $2.15 a day and 82% live in rural areas (19).

The intestinal parasitic infections that are endemic to Malawi include STH and *Schistosoma mansoni* (20). Treatment delivered to school children, using albendazole against STH and praziquantel against schistosomiasis, was scaled up nationwide in 2012 and still continues once annually (17). MDA against onchocerciasis, in the form of ivermectin, is also being conducted annually in eight southern districts.

### Data sources

Data on *A. lumbricoides*, *T. trichiura* and hookworm (*Necator americanus*, *Ancylostoma duodenale*) prevalence, obtained from surveys in schools from 1998 to 2018 in Malawi, was downloaded from ESPEN (21). ESPEN collates data on NTDs in Africa, shared by national governments, and summarises progress to elimination. Prevalence of STH species from 2012 to 2019 was also obtained separately from the Malawi national STH control programme. Lymphatic filariasis MDA programme data, including coverage and the number of rounds completed by each district was obtained from the LF national control programme in Malawi.

Environmental variables that are known to potentially correlate with STH prevalence (22) were selected and downloaded from open sources. An eight day average of Land Surface Temperature (LST) with a 1km resolution and a 16 day composite of the Enhanced Vegetation Index (EVI) at 250m resolution were obtained from the Terra MODIS version 6.0 products (23, 24). A monthly total of precipitation at 0.05 degree resolution was obtained from the Climate Hazards Group InfraRed Precipitation with Station Data (CHIRPS) project (25). Elevation at 30m resolution was obtained from the Shuttle Radar Topography Mission (SRTM) (26). Measures of population size at a 100m resolution and the proportion of individuals who lived under $2 or $1.25 a day from 2010-2011 at 1km resolution were downloaded from the WorldPop database (27). The percentage of households with access to various water, sanitation and hygiene (WASH) indicators were obtained from predictions based on a Bayesian geospatial model at a 5 km resolution previously published (28).

R software version 4.2.2 was used to combine and clean STH prevalence data as well as organise variables for assessment as risk factors for STH infection. All mapping was done on R or QGIS version 3.16. First, any differences between the ESPEN and programme prevalence data were investigated using the geographical coordinates of the survey sites. All survey sites from the programme prevalence data were present in the ESPEN data. However, the ESPEN data had nine sites surveyed in 2017 that were not apparent in the programme data. Therefore, the prevalence from 2019 in the programme data not available through ESPEN, was combined with the ESPEN data and that was taken forward for further analysis.

### Exploratory analysis

*A. lumbricoides*, *T. trichiura* and hookworm prevalence were assessed separately and taken forward for exploratory analysis if there was considerable variation in the observed prevalence. The proportion of data available in each time period, before, during and after LF MDA, was also assessed and a time period was excluded if it constituted less than 5% of the total data. The variables selected were done so using knowledge of factors associated with STH transmission obtained from previous publications and research. These included land surface temperature, monthly precipitation, elevation, the enhanced vegetation index (EVI), the percentage of individuals with access to unimproved sanitation, the percentage of individuals living under $2 a day and the population count. Plots of the variables against the empirical logit transformation (29) of either *A. lumbricoides* or hookworm prevalence were generated to visually identify non-linear relationships. Continuous variables were also standardised. Next, univariate analyses with a logistic mixed effects regression model were conducted to select the best type of summary measure, e.g. mean or maximum precipitation, of relevant variables based on the Akaike Information Criterion (AIC), the lowest AIC indicating a better fit. In other cases, one variable was selected to represent a type of factor, e.g. unimproved sanitation access to represent WASH, based on the AIC and its relative impact on STH transmission.

### Variogram for assessing spatial correlation

The following Bayesian mixed-effects logistic regression model was fitted using the rstanarm package (30). Weakly informative prior distributions, a normal distribution with mean 0 and variance 2.5, were used for the intercept and the coefficients. A variogram was then created to assess the presence of residual spatial correlation.

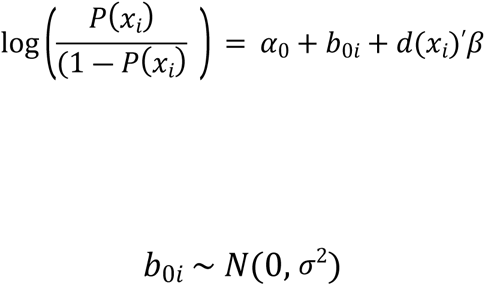

Here the prevalence of either *A. lumbricoides* or hookworm at school *x*_*i*_ is equal to the global intercept (*α*_0_), plus the intercept for school *x*_*i*_(*b*_0*i*_) and d(*x*_*i*_)′*β*, which is the vector of explanatory variables at school *x*_*i*_ with *β* as their coefficients. *b*_0*i*_ at each school are a set of independent zero-mean Gaussian variables with variance *σ*^2^. Estimates of *b*_0*i*_ are used to create the variogram, which was done using the R package geoR (31). Evidence of spatial correlation is determined when the computed variogram lies outside of the 95% confidence intervals (CI), which are computed by 10,000 random combinations of *b*_0*i*_ estimates amongst the sampled schools, assuming spatial independence (32).

### Non-spatial Bayesian model

In the case of no evidence for spatial correlation, the previous model was taken forward and Markov chain Monte Carlo (MCMC) simulations were run to estimate model parameters (33). A total of 10,000 simulations over four chains were run to obtain the resulting estimates and their posterior distributions. The posterior distribution of the parameters were summarised, with odds ratios presented alongside 95% credible intervals. The posterior predictive distribution, which includes the model predictions for the prevalence at schools surveyed in the dataset, was investigated and compared to the actual observed prevalence to assess model fit. Predictions of either *A. lumbricoides* or hookworm prevalence across Malawi, after the termination of LF MDA, were produced from this model on a 2 km^2^ grid alongside their 95% credible intervals. These were then used to determine and map the probability of locations exceeding the thresholds of 2% and 20% *A. lumbricoides* and hookworm prevalence respectively.

### Sensitivity analysis

As additional districts were surveyed after LF MDA, the *A. lumbricoides* and hookworm models were run again with only the districts surveyed both during and after LF MDA. This was done to identify whether the impact of terminating LF MDA on the odds of infection were maintained.

## Results

### Data characteristics and observed prevalence

A total number of 1076 schools were surveyed between 1998 and 2019 across the 28 districts of Malawi. There were a total of nine schools where coordinates were not available or incorrect, reducing the dataset to 1067. At 40 school locations, data on WASH variables were not available, making the final dataset used 1027. Majority of schools sampled around 30 school children across the years with 29% sampling more. The majority of the data, 64%, was collected after LF MDA (2015–2019). Before LF MDA, only 33 schools were surveyed nationwide, averaging around one school per district.

LF MDA coverage in Malawi achieved above 65% coverage for over five years, fulfilling the target for LF MDA. It was implemented between 2008 and 2014 and the average LF MDA coverage nationwide ranged from 73% to 84% across the years. Two districts were classed as non-endemic for LF, Chitipa and Likoma Island. Chitipa received one round of LF MDA in 2014. Likoma received two rounds, in 2010 and 2014 respectively. Of the STH species, *T. trichiura* was the least prevalent in the country, averaging 0.2% (range: 0-1.8%), 0.1% (range: 0-6.7%) and 0.2% (range: 0-7.1%) before, during and after LF MDA respectively. *A. lumbricoides* was most prevalent in school children after the termination of LF MDA, with a national average of 2.5% (range: 0-15.4%), 1% (range: 0-16.7%) and 2.4% prevalence (range: 0-36.7%) before, during and after LF MDA respectively, with hotspots in southern and northern districts (Fig 1). The mean hookworm prevalence nationwide during the three LF MDA phases were 9.8% (range: 0-77.3%), 2.3% (range: 0-66.7%) and 1.4% (range: 0-23.3%), with hotspots in central and northern districts (Fig 2). After LF MDA, variability in the observed prevalence of *A. lumbricoides* increased (Fig 3). This is in contrast with hookworm prevalence, where variability decreased post LF MDA (Fig 3).

**Fig 1.**
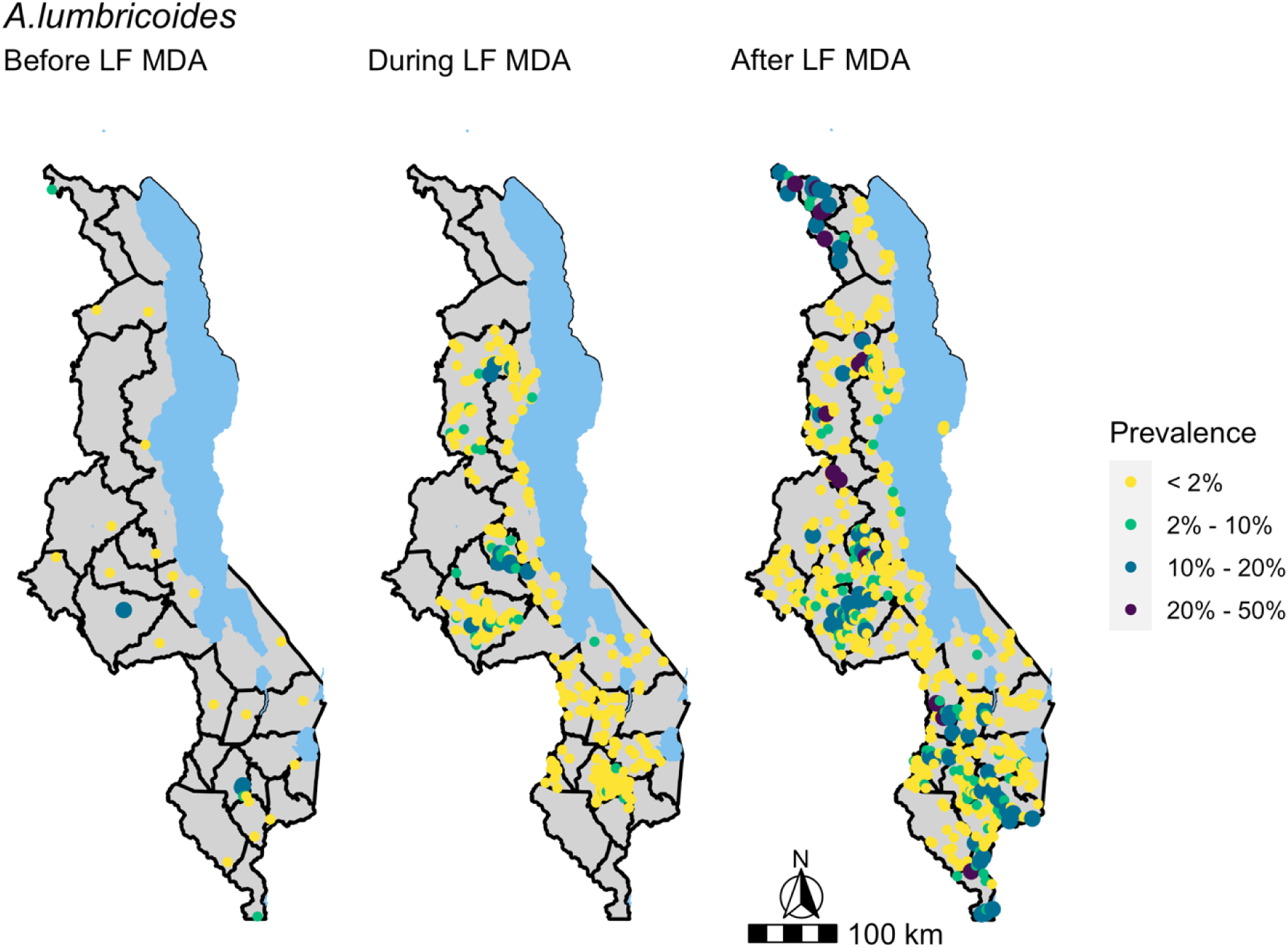
*A. lumbricoides* prevalence in school-aged children during the three phases of LF MDA, before, during and after. Data was obtained from ESPEN and the National Schistosomiasis & STH control programme in Malawi. Survey years range from 1998 to 2019. 33 schools were surveyed before LF MDA (1998–2004), 337 schools during LF MDA (2012–2014) and 657 after LF MDA (2015–2019).

**Fig 2.**
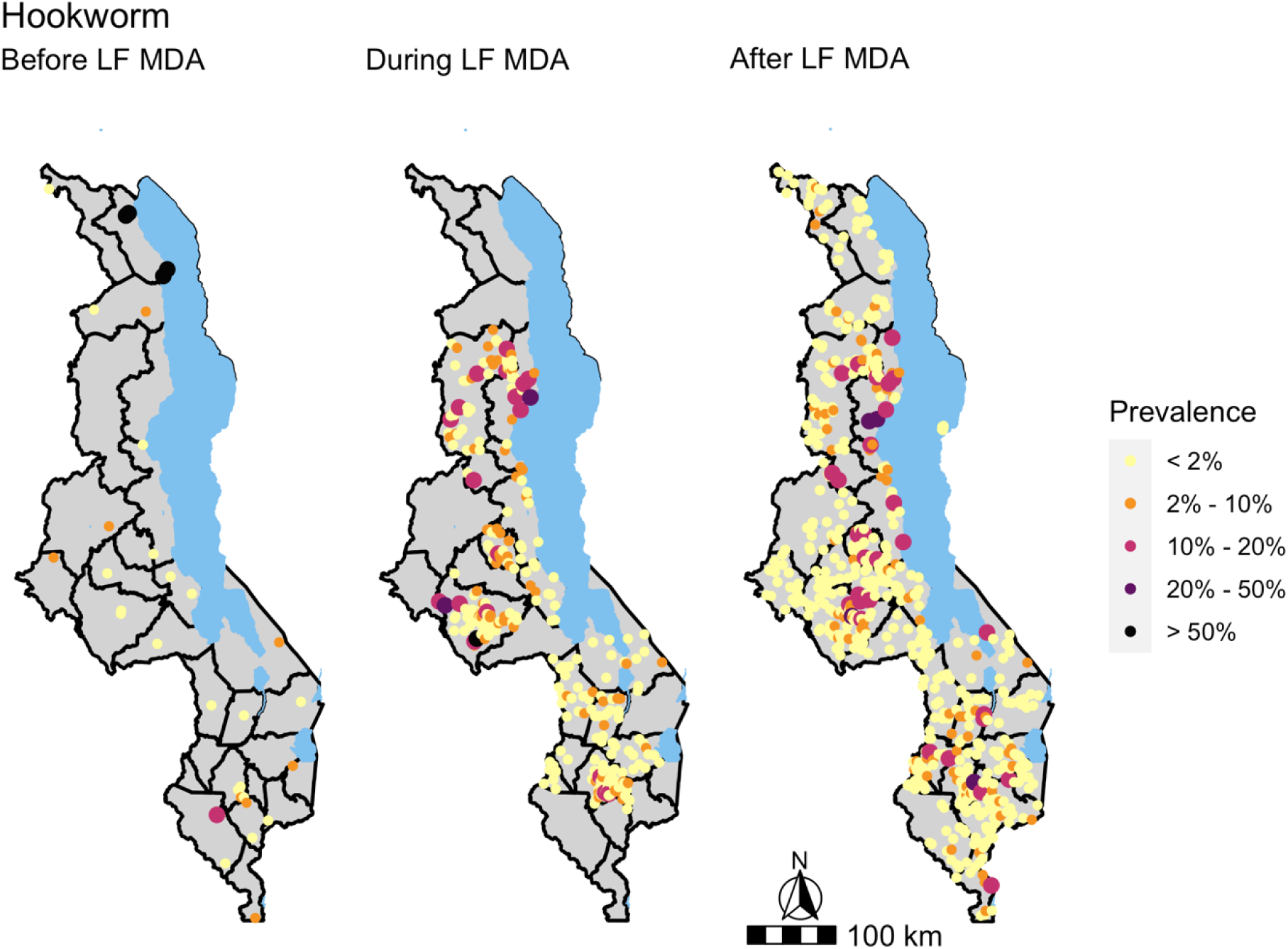
Hookworm prevalence in school-aged children from during the three phases of LF MDA, before, during and after. Data was obtained from ESPEN and the National Schistosomiasis & STH control programme in Malawi. Survey years range from 1998 to 2019. 33 schools were surveyed before LF MDA (1998–2004), 337 schools during LF MDA (2012–2014) and 657 after LF MDA (2015–2019).

**Fig 3.**
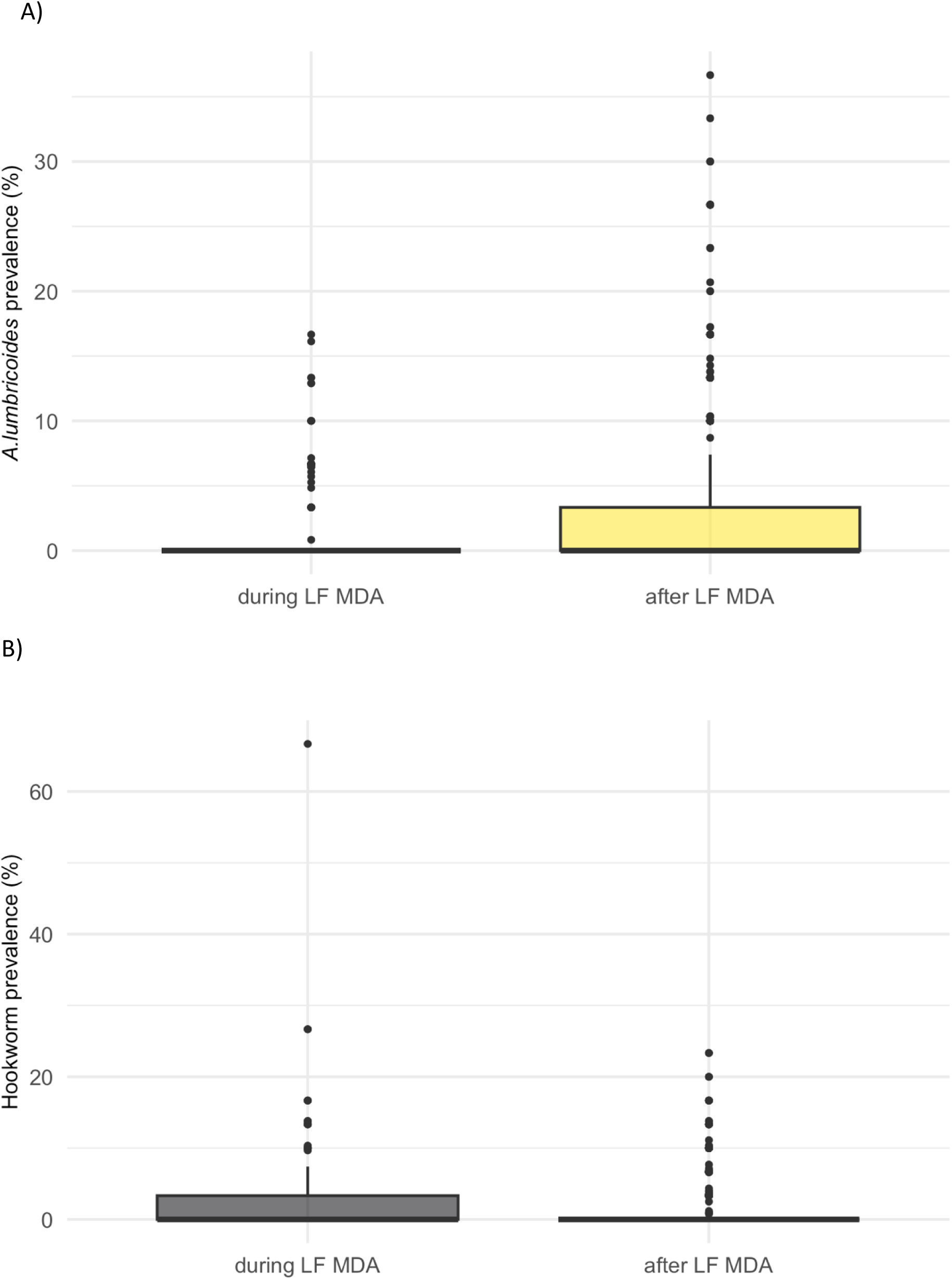
Prevalence in school aged children during and after LF MDA in Malawi. (A) *A. lumbricoides*. (B) Hookworm. Data was obtained from ESPEN and the National Schistosomiasis & STH control programme in Malawi. 337 schools were surveyed during LF MDA (2012–2014), 657 schools surveyed after LF MDA (2015–2019).

Due to the sparsity of data before LF MDA, which only had 33 schools in total, these points were excluded from further modelling. A model was also not fit to *T. trichiura* as its prevalence range only extended to 7% prevalence.

### Assessment of spatial correlation

Variograms for assessing the presence of spatial correlation are shown in Fig 4. The variograms did not exhibit the distinct pattern of an initial increase in variance between schools with increasing distance and the eventual plateau as the variance reaches its maximum, suggesting the absence of spatial correlation. Instead, variance between schools fluctuates around the same level regardless of distance with both species.

**Fig 4.**
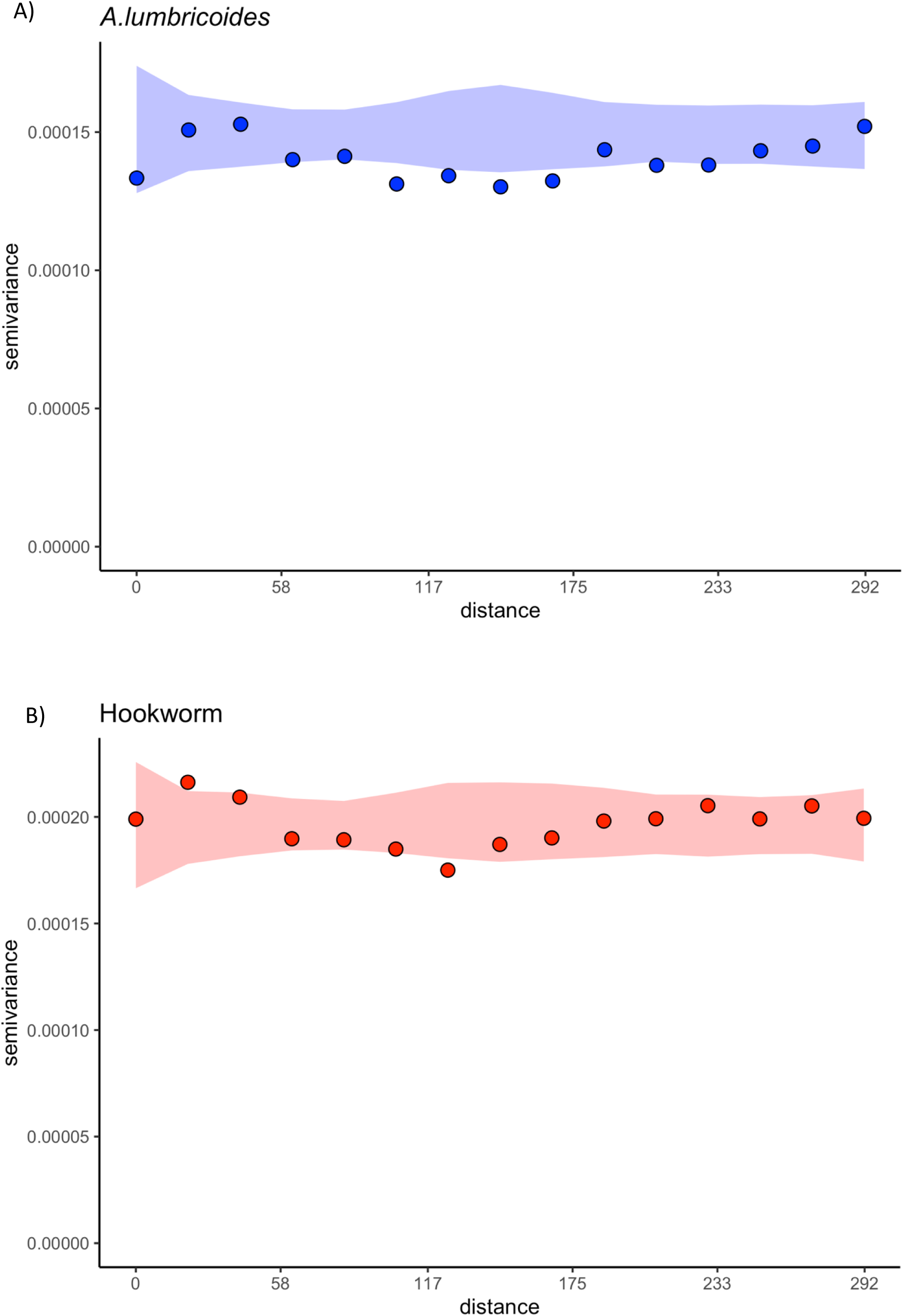
Variograms to assess the presence of residual spatial correlation with distance between schools on the x axis, variance on the y axis and the shaded areas the 95% confidence intervals. (A) *A. lumbricoides.* (B) Hookworm.

### Determinants of *A. lumbricoides* and Hookworm infection

The results from the mixed-effects Bayesian logistic regression, accounting for school level heterogeneity, is shown in Fig 5. School children after the termination of LF MDA had a three and a half time increase in the odds of *A. lumbricoides* infection compared to school children during the implementation of LF MDA (Odds Ratio (OR): 3.4, 95% credible interval (CI): 1.99 – 5.94). In contrast, odds of hookworm infection were lower after LF MDA (OR: 0.5, 95% CI: 0.33 – 0.73). Increase in precipitation in the previous year was associated with both increased odds of *A. lumbricoides* (OR: 2.16, 95% CI: 1.47 – 3.23) and hookworm infections (OR: 1.56, 95% CI: 1.16 – 2.1). Higher elevations were associated with increased odds of *A. lumbricoides* infection (OR: 1.66, 95% CI: 1.26 – 2.22). An increased level of EVI and larger populations were associated with increased odds of hookworm infection, with odds ratios of 1.44 (95% CI: 1.1 – 1.9) and 1.23 (95% CI: 1.02 – 1.49) respectively. The variability between the schools, as indicated by the school standard deviation obtained from the models, can be compared before and after the inclusion of the explanatory variables in the models presented. The variability between the schools after addition of the explanatory variables was only reduced by 5.7% and 4% in the *A. lumbricoides* and hookworm models respectively (S1 Table).

**Fig 5.**
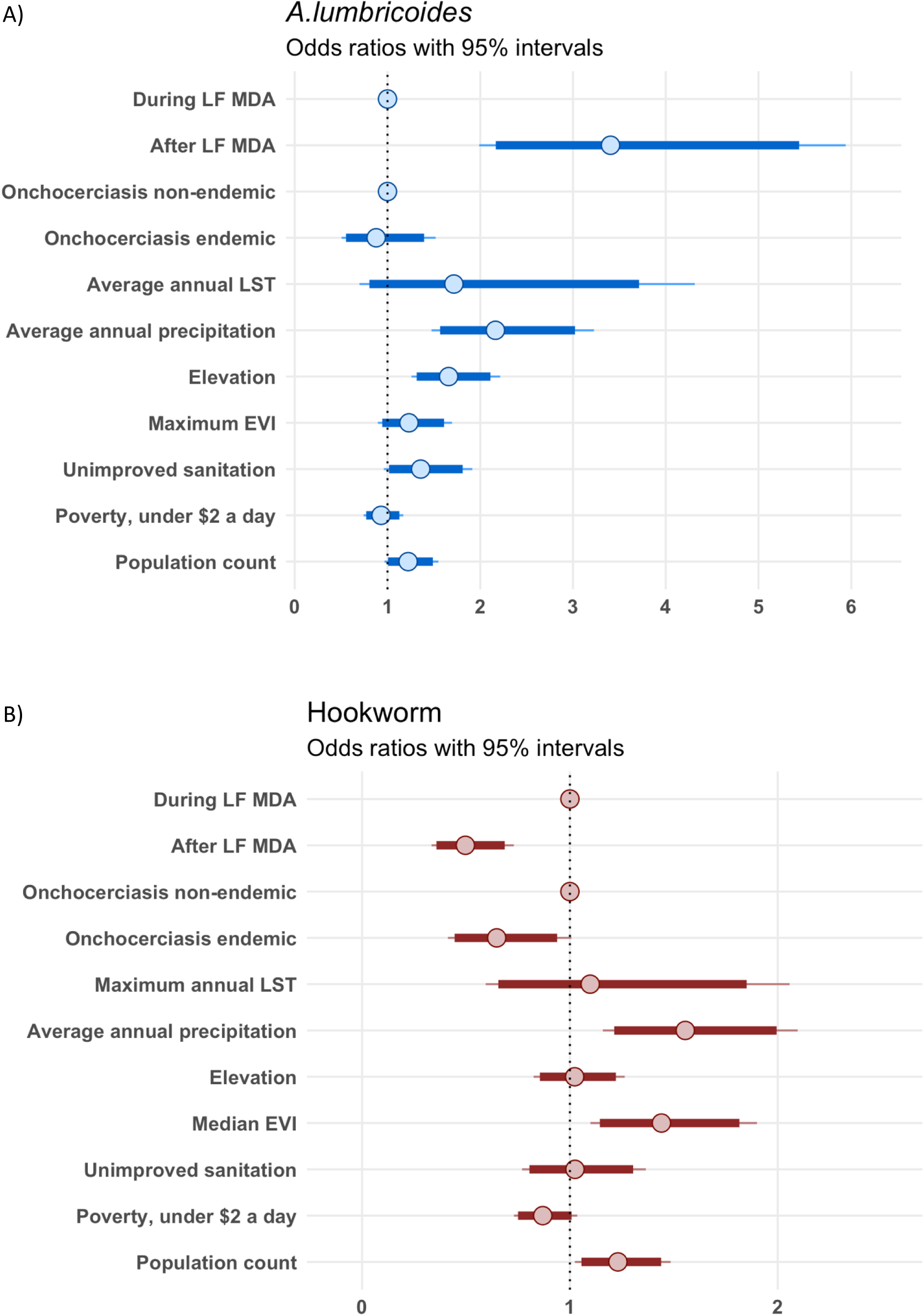
Determinants of *A. lumbricoides* (A) and hookworm (B) prevalence in school aged children in Malawi, obtained from a multivariate mixed-effects Bayesian logistic regression model.

### Assessment of model fit

The posterior predictive distributions at schools surveyed are shown in Fig 6. To assess model fit, 10,000 simulations of the prevalence predicted by the model at each school are compared to the observed prevalence at each school. Majority of the predictions generated from the *A. lumbricoides* model showed increased variance in prevalence in comparison to the variance observed in the dataset, captured by the range of standard deviations simulated (Fig 6b). Similarly, the maximum prevalence observed in the dataset is in the lower end of the distribution of the maximum prevalence’s predicted by the model (Fig 6c). The hookworm model showed the same patterns, the model predominantly predicted both a higher maximum hookworm prevalence and exhibited increased variance (Fig 6e, 6f). However, the mean *A. lumbricoides* and hookworm prevalence were well captured by the model, with the observed mean in the middle of the distribution (Fig 6a, 6d).

**Fig 6.**
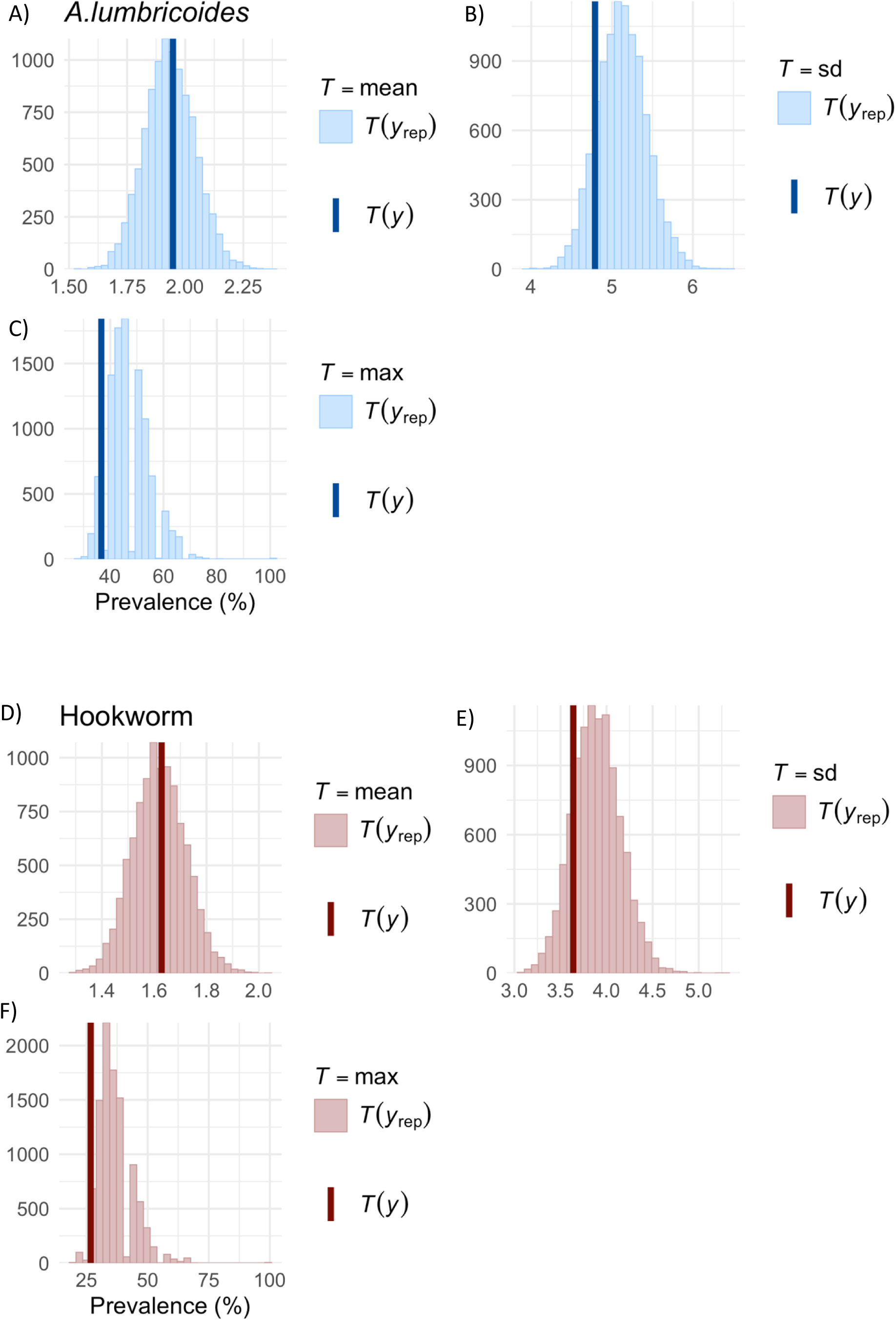
Density plots of summaries of the posterior predictive distribution at surveyed schools, generated from 10,000 simulations of a multivariate mixed-effects Bayesian logistic regression model. (A-C) Density plot of the mean, standard deviation, and maximum prevalence predicted by the *A. lumbricoides* model for each of the 10,000 simulations. (D-F) Density plot of the mean, standard deviation and maximum prevalence predicted by the hookworm model for each of the 10,000 simulations. *y*_rep_ = simulations, *y* = observed.

### Predictions

Nationwide predictions of *A. lumbricoides* and hookworm prevalence in school children post LF MDA were generated from the models presented. The raw predictions with 95% credible intervals are available in S2 Fig and S3 Fig. For both species, the credible intervals have a wide range, where it extends from 0% prevalence nationwide at the lower limit to above 50% predicted prevalence, in selected areas, at its upper limit (S2 Fig, S3 Fig). The probability of exceeding either 2% or 20% prevalence are shown in Fig 7 and Fig 8. For *A. lumbricoides* infections, there are patches where the probability of exceeding 2% prevalence is above 50%. These include some districts in the south, in particular Mulanje district, at borders between northern districts and areas in central Malawi. These same areas have between 10% and 30% probability of exceeding 20% prevalence with the exception of Mulanje district, where the probability is above 50%. Areas that have low elevation in the south have a less than 20% probability of exceeding 2% *A. lumbricoides* prevalence. Map of elevation is available in S4 Fig. For the predictions of hookworm prevalence, there are fewer areas where the probability of exceeding 2% prevalence is above 50%. These are in the north just west of Lake Malawi, in Mulanje district and at borders in northern districts. These areas also have between 10 to 30% chance of exceeding 20% hookworm prevalence.

**Fig 7.**
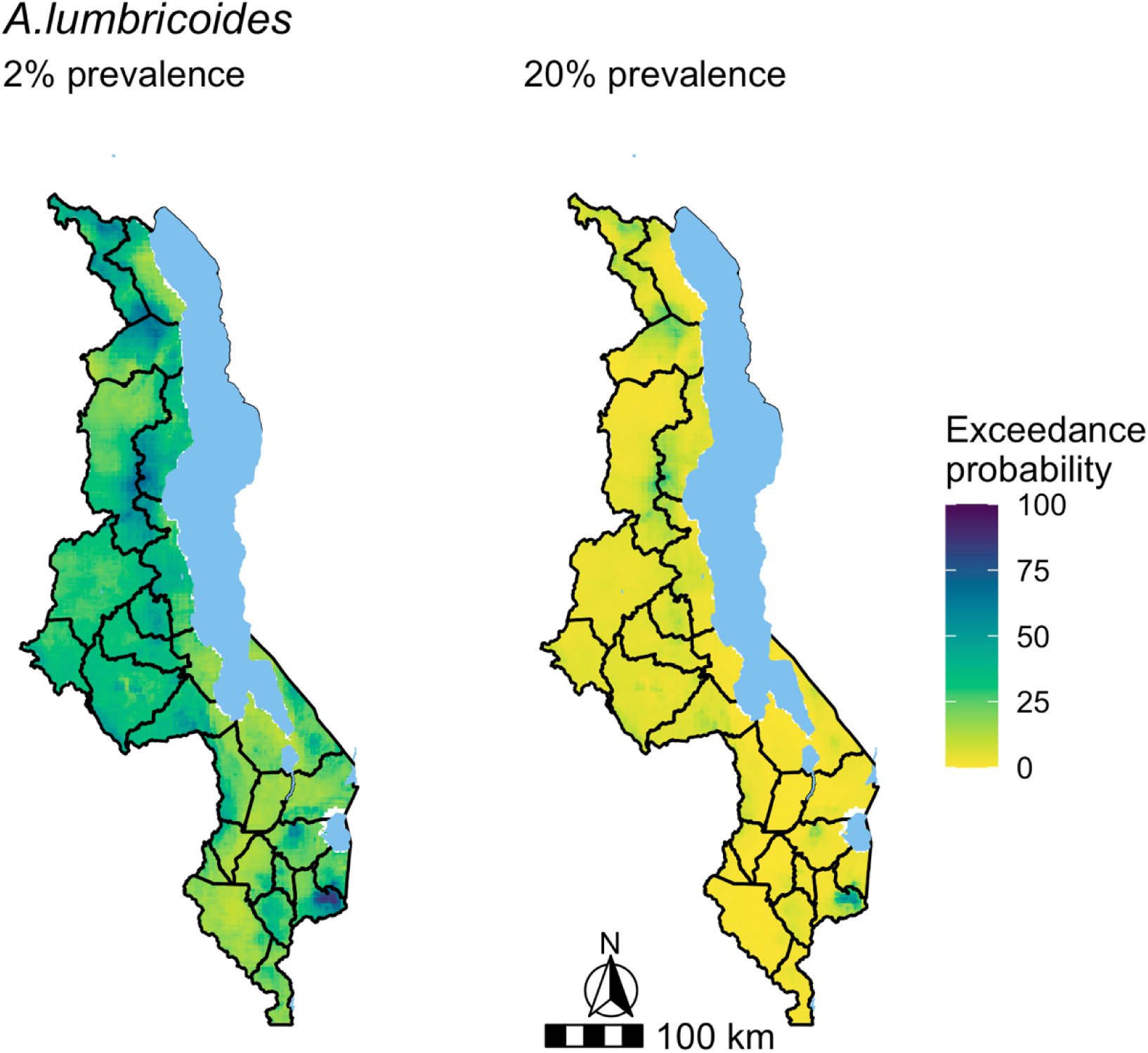
Maps of the probability of exceeding 2% or 20% predicted *A. lumbricoides* prevalence in Malawi. Predictions were generated from a multivariate mixed-effects Bayesian logistic regression model.

**Fig 8.**
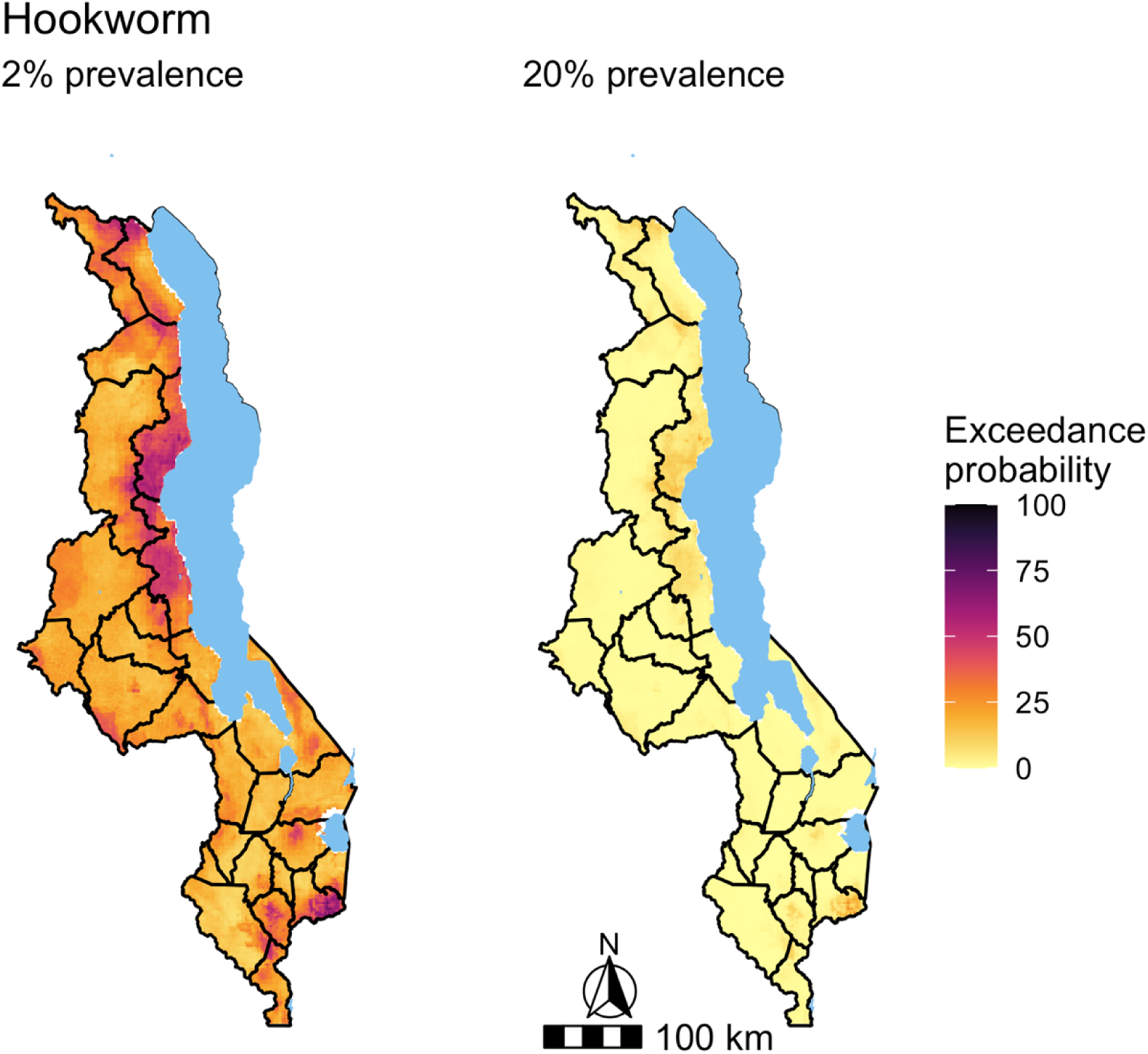
Maps of the probability of exceeding 2% or 20% predicted hookworm prevalence in Malawi. Predictions were generated from a multivariate mixed-effects Bayesian logistic regression model.

In summary, there are only a number of areas where there is an above 50% probability that *A. lumbricoides* prevalence exceeds 2 or 20%. In the case of hookworm prevalence, the number of areas is fewer in regards to exceeding 2% prevalence and the likelihood of exceeding 20% prevalence is below 50% probability nationwide. However, there is an overlap in the areas highlighted between the species. These also correlate to regions that receive more rainfall and are of higher elevation (S4 Fig).

### Sensitivity analysis

Fig 9 presents the output of the sensitivity analysis where the models were restricted to only include districts surveyed in both time periods, during and after LF MDA. This reduced the total number of schools from 994 to 753, 337 surveyed during and 416 surveyed after LF MDA. The *A. lumbricoides* model, continues to indicate increased odds of infection in school children post termination of LF MDA (OR: 2.97, 95% CI: 1.58 – 5.72) and the presence of a resurgence in infection. With the hookworm model, odds of infection post LF MDA are lower when compared to during LF MDA, but this is no longer significant. Areas endemic for onchocerciasis are now significantly associated with lower odds of hookworm infection.

**Fig 9.**
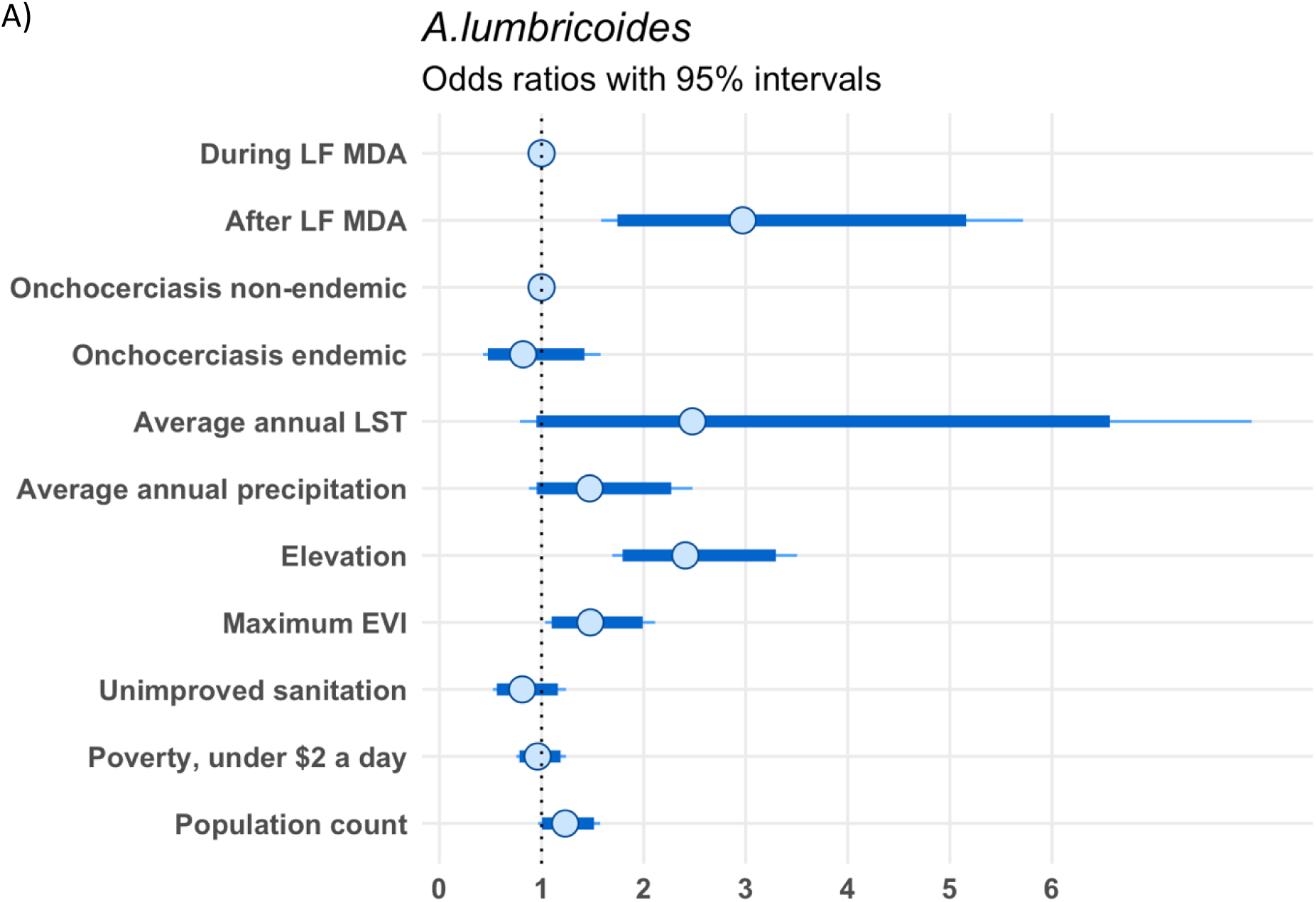
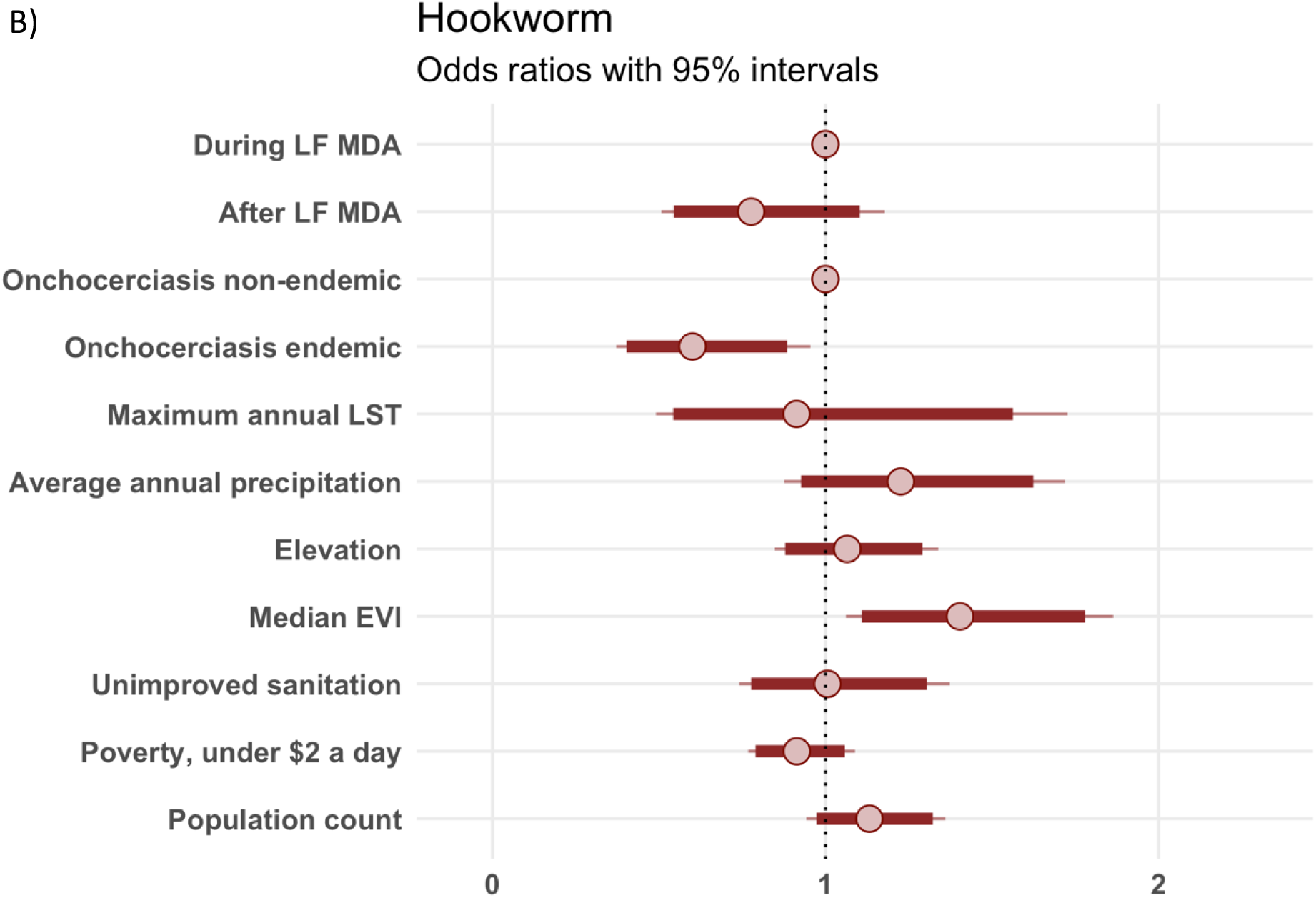
Impact of LF MDA termination on *A. lumbricoides* (A) and hookworm (B) prevalence in school aged children in Malawi.

## Discussion

This analysis of STH prevalence data from Malawi reveals a resurgence in *A. lumbricoides* infections in school children post implementation of LF MDA, despite ongoing annual STH PC targeting school aged children.

The overall, country-wide odds of infection with *A. lumbricoides* were found to have increased as much as three-fold after the termination of LF MDA, and this effect was maintained when conducting the sensitivity analysis, which limited the analysis to districts sampled both during and after LF MDA. In contrast to *A. lumbricoides*, our hookworm model suggests that hookworm infection risk significantly decreased after several years of community wide LF MDA, but this result was not confirmed by the sensitivity analysis.

Adults are estimated to host around 30% of the infectious reservoir of *A. lumbricoides*, which could partially explain this resurgence (34). It is conceivable that in Malawi, the community wide treatment set *A. lumbricoides* prevalence to a low transmission level and reduced infectious eggs in the environment, which could not be maintained once community treatment was stopped. A previous study also showed that *A. lumbricoides* has a quicker re-infection rate than hookworm, where around 15% of individuals infected with *A. lumbricoides* and treated with albendazole, were found infected 8-9 months after. In the case of hookworm, this value was only 3% (35). Additionally, hookworm infection levels are known to be lower in children than in adults, with infections peaking at around middle age to 60 years old (5). This could explain why prolonged community wide LF MDA and continuing STH PC may have decreased hookworm infection levels in school children enough to be undetectable by routine monitoring surveys. However, there is the issue of diagnostic accuracy. The Kato-Katz technique used in the monitoring approach lacks sensitivity, preventing the detection of light infections that are expected to prevail in areas of historic or ongoing community wide distribution of drugs efficacious against STH (36, 37). This issue is particularly salient for hookworm due to the additional challenge of egg frailty, which are rapidly destroyed following Kato Katz slide preparation (36, 38, 39). This is likely a major influence in the low hookworm levels detected post LF MDA.

The *A. lumbricoides* model predictions indicate areas in the extreme south of the country as likely “hotspots”. However, the predictions had wide credible intervals and primarily follow the distribution of precipitation and elevation in Malawi, as these were the two factors positively associated with infection risk in our *A. lumbricoides* model. These associations are well documented in many settings and is in line with biological plausibility, as survival of STH is favoured in moist and warm environments as well as at higher altitudes, where heat, which can cause egg desiccation, is less extreme (4, 40–44).

In terms of human-activity related determinants, we found a borderline significant trend of a positive association between *A. lumbricoides* infection risk and both increasing unimproved sanitation coverage and population. With hookworm, odds of infection were significantly higher in areas with larger populations. The association between WASH and STH infection levels and transmission is widely acknowledged (45–48). Larger populations with poor sanitation infrastructure can increase transmission due to higher environmental contamination (49, 50). However, inconsistency in finding associations with WASH and socioeconomic factors have been noted, citing high heterogeneity between studies and the lack of intervention trials assessing these factors (51–53).

### Limitations

There are limitations to this analysis.

First, the models could not be adjusted for STH PC coverage, due to data unavailability before 2016. Assumptions were not made as reports from the national schistosomiasis and STH control programme in Malawi describe heterogeneity in albendazole availability for STH PC across different years, making assumptions of coverage more difficult (17).

Another limitation is the lack of infection intensity data. A community based cross-sectional study from 2018 in Mangochi district found all STH infections were of light intensity (54), which matches findings from other post LF MDA settings (13–16). The absence of this information limits result interpretation in terms of public health impact and prevents assessing whether the WHO target for morbidity control of < 2% prevalence of M&HI infections has been met (6).

Predictions of both *A. lumbricoides* and hookworm infection showed uncertainty, reflected by the wide credible intervals of the predictions. This uncertainty as well as the lack of reduction in the variability between schools in the data after the explanatory variables were included in the models, suggests that our models were lacking important predictive factors for *A. lumbricoides* and hookworm infection. These could include higher resolution sanitation coverage data or extreme weather events which could have impacted transmission. The role of human population movement or the potential emergence of drug resistance should also considered.

Movement of people into an infected area or their movement bringing parasites into a previously uninfected area, is a key factor in the epidemiology of infectious diseases (55–57).

A model simulation has shown that as few as 2% of the population of an area, even one of low prevalence, moving to an area that has undergone community based treatment for five years, is enough to reduce the probability of eliminating both *A. lumbricoides* and hookworm in the treated area to below 50% (58). The potential of reduced drug efficacy through either genetic resistance or other mechanisms, should also be investigated when assessing trends in STH transmission and intervention impact In Malawi, especially when MDA against onchocerciasis, LF and STH PC have been conducted for more than two decades (59).

### STH PC in other contexts

The increase in *A. lumbricoides* prevalence after the termination of LF MDA suggests that the benefits of a chemotherapy-based intervention as intensive as LF MDA cannot be sustained through STH PC only targeting school aged children. Moreover, this quick rise in *A. lumbricoides* prevalence after 6 years of LF MDA at around 78% coverage, also support findings from modelling studies suggesting that community wide treatment is needed to reach the WHO morbidity target when compared to school based distributions (10, 11).

Examples of STH PC in other countries also indicate this. In Angola, after 6 years of STH PC, none of the provinces investigated showed a significant reduction in STH prevalence nor achieved the target of < 2% of M&HI infections, with one province showing a particularly high prevalence of 25% M&HI infections (60). In Kenya, significant reductions from baseline across all species were seen after 5 years of PC but certain regions still displayed STH prevalence above the WHO threshold of 20% (61).

### Conclusion

A major obstacle to accurately monitor STH infection levels in the era of large-scale chemotherapy-based interventions is the low sensitivity of currently available methods, including those used to generate the data in this study. The actual resurgence in *A. lumbricoides* infection in Malawi is therefore likely to be higher than the observed threefold increase in the odds of infection. A study conducted in a district in Mozambique showed the impact of low sensitivity on predicting prevalence. In a low infection intensity setting, with 5 years of STH PC, the use of qPCR diagnosis resulted in 86% of neighbourhoods predicted to have a prevalence above 20% when modelling the data. This is compared to just 10% of neighbourhoods estimated to have 20% prevalence or more when diagnosing using a single Kato-Katz (62).

The need for high sensitivity diagnostics is widely acknowledged by the STH scientific and public health community. Only such diagnostic tools will allow accurately estimating infection levels, intervention impact and better delineate infection hotspots in which monitoring should be prioritised. A greater assessment of underlying factors would increase understanding of current STH transmission and further aid the identification of hotspots. In the meantime, surveys using more intensive diagnostic approaches such as multiple slides on multiple samples could be conducted in areas of concern.

Monitoring activities are urgently required in Malawi to better understand infection transmission, factors underlying the potential resurgence in *A. lumbricoides* infections and how this translates to infections of M&HI, which is the focal point of the WHO targets for STH control.

## Data Availability

The data that support the findings of this study are publicly available from the Expanded Special Project for Elimination of Neglected Tropical Diseases at https://espen.afro.who.int/. A portion of data was also obtained and available from the STH national control programme and LF national control programme in Malawi.

